# The science of child and adolescent mental health in Brazil: a nationwide systematic review and compendium of evidence-based resources

**DOI:** 10.1101/2024.11.10.24317061

**Authors:** Lauro Estivalete Marchionatti, André Cardoso Campello, Jessica Azevedo Veronesi, Carolina Ziebold, André Comiran Tonon, Caio Borba Casella, Julia Luiza Schafer, Aaliyah Nadirah Madyun, Arthur Caye, Christian Kieling, Luis Augusto Rohde, Guilherme V. Polanczyk, Jair Mari, Rudi Rocha, Leonardo Rosa, Dayana Rosa, Zila M Sanchez, Rodrigo A Bressan, Shekhar Saxena, Sara Evans-Lacko, Pim Cuijpers, Kathleen R. Merikangas, Brandon A. Kohrt, Jason Bantjes, Shirley Reynolds, Zeina Mneimneh, Giovanni Abrahão Salum

## Abstract

**Background:** Brazil is home to 50 million children and adolescents, whose mental health needs require context-sensitive research. Although scientific output is growing in the country, publications are scattered and often inaccessible.

**Methods:** This systematic review compiles prevalence estimates, assessment instruments, and interventions for child and adolescent mental health in Brazil (PROSPERO registration: CRD42023491393). We searched international (PubMed, Web of Science, PsycINFO, Google Scholar) and national (Scielo, Lilacs, Brazilian Digital Library of Theses and Dissertations) databases up to July 2024. Reference lists, reviews, and experts were consulted. Extraction followed Consensus-based Standards for the selection of health Measurement Instruments (COSMIN) and Cochrane manuals.

**Results:** This review appraises 734 studies on 2,576 prevalence estimates, 908 studies on 912 instruments, and 192 studies on 173 intervention trials. Point prevalence of any mental disorder ranged from 10.8% (age 12) to 19.9% (ages 7 to 14), although a nationally-representative study is lacking. There is a rise of self-harm notifications, reaching 133.1 in 2019 (per 100,000 aged 10-19). Indigenous youth face suicide rates of 11 (age 10 to 14), far exceeding national averages (0.652). There is severe violence exposure (21% of adolescents reported domestic physical violence in the previous month), disproportionately impacting Black youth and heightening risks for depression and substance use. Reliable instruments exist for assessing psychopathology, yet most lack psychometric and cross-cultural validation. Interventions remain under-implemented; the largest trials adapted international substance-use prevention programs, showing null effects. High-quality studies are mainly funded by public investment.

**Discussion:** This compilation provides accessible data for professionals, facilitating translation of science to practice. Brazilian sociocultural challenges impact youth mental health, with public health priorities including violence, systemic racism, and indigenous youth suicide. National research must develop culturally-sensitive resources for mental health, including scalable interventions focused on social minorities.

**Funding:** The Stavros Niarchos Foundation.

## Introduction

Brazil, the largest country in the Global South, is home to 50 million children and adolescents, yet this population has been historically neglected by the country’s public policies.^1,2^ Despite early 21^st^-century growth, the country grapples with severe inequality, high poverty and violence rates, and economic downturns.^3,4^ Brazilian youth experience adverse environments, with estimations that approximately 20% of those aged 10 to 19 suffer from a mental disorder.^5-7^ The Psychosocial Care Network (“*Rede de Atenção Psicossocial”*, or *RAPS*) addresses this needs, operating as a community-based mental health system within the Brazil’s Unified Health System (“*Sistema Único de Saúde”*, or *SUS*).^8^ Despite notable achievements, *RAPS* is strained by underfunding, coverage gaps, and quality deficits. The educational system is also critical for mental health, with the 2024 National Policy for Psychosocial Care in School Communities established to address shortages in programs, resources, and specialized staff in public schools.^9^

Mental health practices are inconsistent across the country, with insufficient uptake of clinical guidelines nationally established for the public system.^8^ While oriented by international literature and suitably determining pharmacological approaches, these protocols are arguably insufficient to address Brazil’s diverse contexts. There is a need for locally-established science and decolonial approaches addressing the unique struggles of Brazilian children and adolescents, including members of indigenous populations, quilombos (communities with a strong African-Brazilian heritage that were originally composed of settlements formed by self-liberated enslaved people), favela residents, and victims of systemic racism.^10-15^

Brazilian science has witnessed significant progress driven by public funding and technological development.^16-18^ Since the 2000s, the country has experienced a surge in scientific publications, growing at more than double the international rate.^19,20^ This has positioned Brazil as the 13^th^ largest global scientific producer, with a notable emphasis on the health field.^21-23^ However, impact metrics have not kept pace; citation indexes remain below global and Latin American averages, and many works go uncited.^21,24^ The scattered nature of publications highlights weaknesses in scientific communication, lacking cross-referencing essential for advancing knowledge.^25,26^ In isolation, articles become costly to locate and appraise, creating an ubiquitous barrier to the uptake of science in practice.

This systematic review aims to identify and appraise prevalence surveys, assessment instruments, and interventions for child and adolescent mental health in Brazil. Our goal is to create an accessible resource for consulting the evidence base, evaluate the current state of research in the field, and take practical insights for policy.

## Methods

We initially conducted an umbrella search, screening studies into three areas: prevalence estimates, assessment instruments, and interventions. Then, we carried out a three-arm review with specific methods for each topic. Our approach was based on a similar country-wide systematic review in Greece.^27^ We followed the Preferred Reporting Items for Systematic Reviews and Meta-Analysis (PRISMA) statement (*Supplementary Table 1*) and the PRISMA-COSMIN manual for reporting systematic reviews of measurement instruments.^28,29^ Protocol was registered with PROSPERO [CRD42023491393].

### Search strategy

We employed a comprehensive strategy without restrictions of language from inception to July 18th, 2024. Our query associated terms related to mental health, to children and adolescents, and to Brazil, being adapted to English and/or Portuguese according to databases (see *Supplementary Table 2*).

The multi-step search procedure consisted of: (1) international databases (PubMed, PsycINFO, and Web of Science); (2) regionally-representative databases (SciELO and Lilacs); (3) Google Scholar; and (4) a national repository of theses and dissertations (*BDTD - Biblioteca Digital Brasileira de Teses e Dissertações*).

This was complemented by: (5) identifying review articles during screening and inspecting their reference lists; (6) snowballing inclusions based on relevant studies; and (7) experts consultation. After extraction, we conducted (8) a specific search on identified gaps (see *Supplementary Table 3*).

References were uploaded to *Rayyan*.^30^ Studies retrieved in steps 1 and 2 were automatically deduplicated using EPPI-Reviewer 4.0,^31^ with a second manually-verified deduplication in Rayyan.

### Screening

We conducted primary screening based on title/abstract, categorizing studies into one or more areas: prevalence, instrument, and intervention. Pairs of independent researchers (ACC and CZ; ACT and CBS) performed this initial screening for studies retrieved from the steps 1 and 2. Discrepancies were discussed and resolved by a third reviewer (LEM) when needed. Data sources described in steps 3 and 4 did not allow for exportation and were manually inspected by a single screener (ACC and CZ). In Google Scholar, results were screened until reaching one hundred consecutive studies without novel inclusions. Due to its crawler-based nature, Google Scholar returns overwhelming results and is recommended as a supplementary datasource.^32^

Secondary screening and data extraction were performed independently for each area. These steps were conducted in a single reviewer procedure, with at least 20% of the articles verified by another team member (LEM). This approach was chosen due to the large scope of this review, which prioritized sensitivity over specificity. The team conducted a pilot calibration of inclusion criteria based on at least 10% of the results and held regular discussion meetings.

### Inclusion criteria

We included studies reporting prevalence rates, assessment instruments, or interventions related to child and adolescent mental health outcomes and risk factors (e.g.: includes mental disorders, substance use, suicidality, maltreatment and neglect, bullying, quality of life, wellness, cognitive neurodevelopment, learning difficulties, personality traits, mental health literacy and awareness, and other associated constructs). The eligible population was children/adolescents up to 19 years-old in Brazil (school-based samples with outliers above 19 years old were also eligible), including assessments/interventions involving caregivers/professionals. Research articles, editorial letters with original data, dissertation theses, and book chapters were eligible.

For prevalence studies, we included surveys on community- and school-based samples using structured instruments or epidemiological registers to report prevalence rates or symptom levels.

Instrument studies were included if they developed, translated, or validated assessment instruments (or solely applied the instrument if this was the unique report on that tool).

Intervention studies were included when reporting experimental designs (from pre-post uncontrolled studies to randomized clinical trials (RCTs)) or adaptation/translation of interventions from other settings.

### Exclusion criteria

We excluded multi-country or general population studies that did not disaggregate results between countries or age groups. We excluded studies on neurodevelopmental disorders reporting only motor or phonetic development, on mental health outcomes of caregivers or professionals that do not directly concern children and adolescents (e.g., maternal depression or teacher burnout). Conference abstracts were uneligible.

We excluded prevalence studies on clinical populations (e.g., quality of life among leukemia patients), on populations with very specific determinants (e.g., survivors of traumatic events), reporting the absolute numbers of suicide or violence notifications without populational rates, and datasets with duplicate information (we only included the most comprehensive and/or updated).

Instrument studies were excluded when reporting guides for qualitative interviews, projective tests, or tools assessing quality of life in the context of clinical conditions.

Intervention studies were excluded when reporting case studies or series with fewer than five participants.

### Data extraction and synthesis

#### Prevalence studies

Procedures were based on a widely-cited systematic review on the prevalence of child and adolescent mental health disorders.^33^ Risk of bias was assessed with a validated tool covering external and internal validity, analysis bias, and representativeness.^34^ Each estimate from each study was a separate entry. After extraction, a synthesis table aggregated all estimates per construct.

#### Instrument studies

The Consensus-based Standards for the Selection of Health Measurement Instruments (COSMIN) guidelines oriented the extraction, quality assessment, and psychometric evaluation.^35^ We employed a three-level procedure encompassing extraction (recording data per each instrument at each study), evaluation summary of its psychometric properties, and synthesis (aggregating data for each instrument) (see details in Supplementary Table 4, Supplementary Table 5 and Supplementary Table 6).

#### Intervention studies

Procedures were based on the Cochrane Manual for Systematic Reviews of Interventions.^36^ Methodological quality was assessed with the revised Cochrane tool for randomized trials (RoB 2) and the Joanna Briggs Institute (JBI) tool for non-randomized designs.^37,38^

## Results

We included 734 studies reporting 2576 prevalence estimates, 904 studies reporting 912 unique assessment instruments, and 192 studies reporting 173 trials (see Figure 1 for flowchart; see Supplementary Table 7 for reasons for exclusions). The full dataset can be navigated in Supplementary File 1.

**Figure 1.**
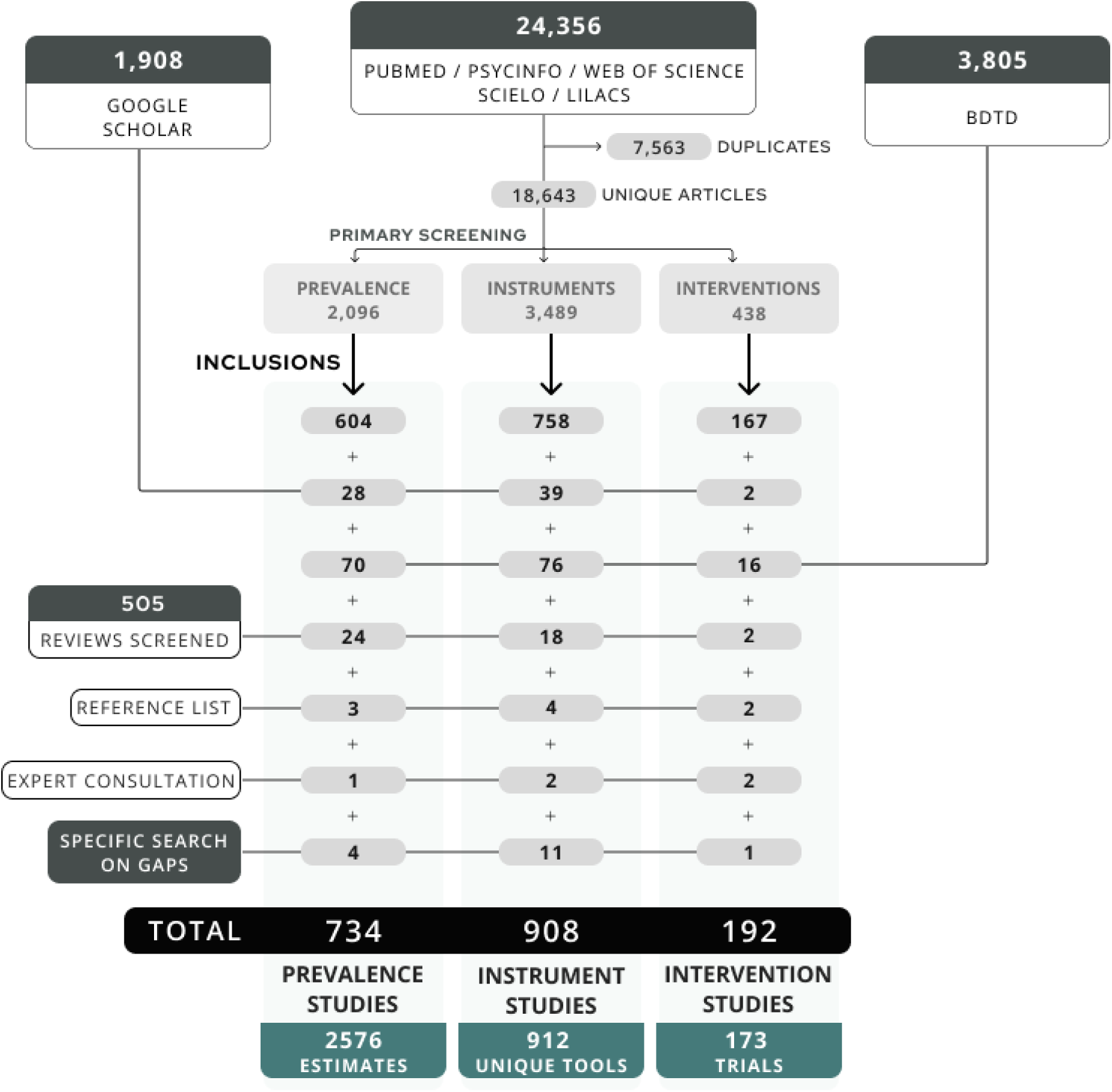
Flowchart describing the search and screening procedures.

### Overview of scientific production

Figure 2 presents a time trend analysis of publications and topics of inquiry (see see heatmap graph in Supplementary Figure 1). Since 2010, Brazilian scientific production on youth mental health has grown significantly, but intervention intervention studies represent only a tenth of all articles. Studies concentrate on topics such as substance use, psychosocial symptoms, exposure to adversities, and depression. There is significantly less focus on areas specifically relevant to children and adolescents, including Autism Spectrum Disorder (ASD), intellectual disability, and learning disorders. A particularly overlooked domain is sexuality and gender. There are only two works on instruments assessing sexual trajectories discrimination,^39,40^ and no reports on gender dysphoria.

**Figure 2.**
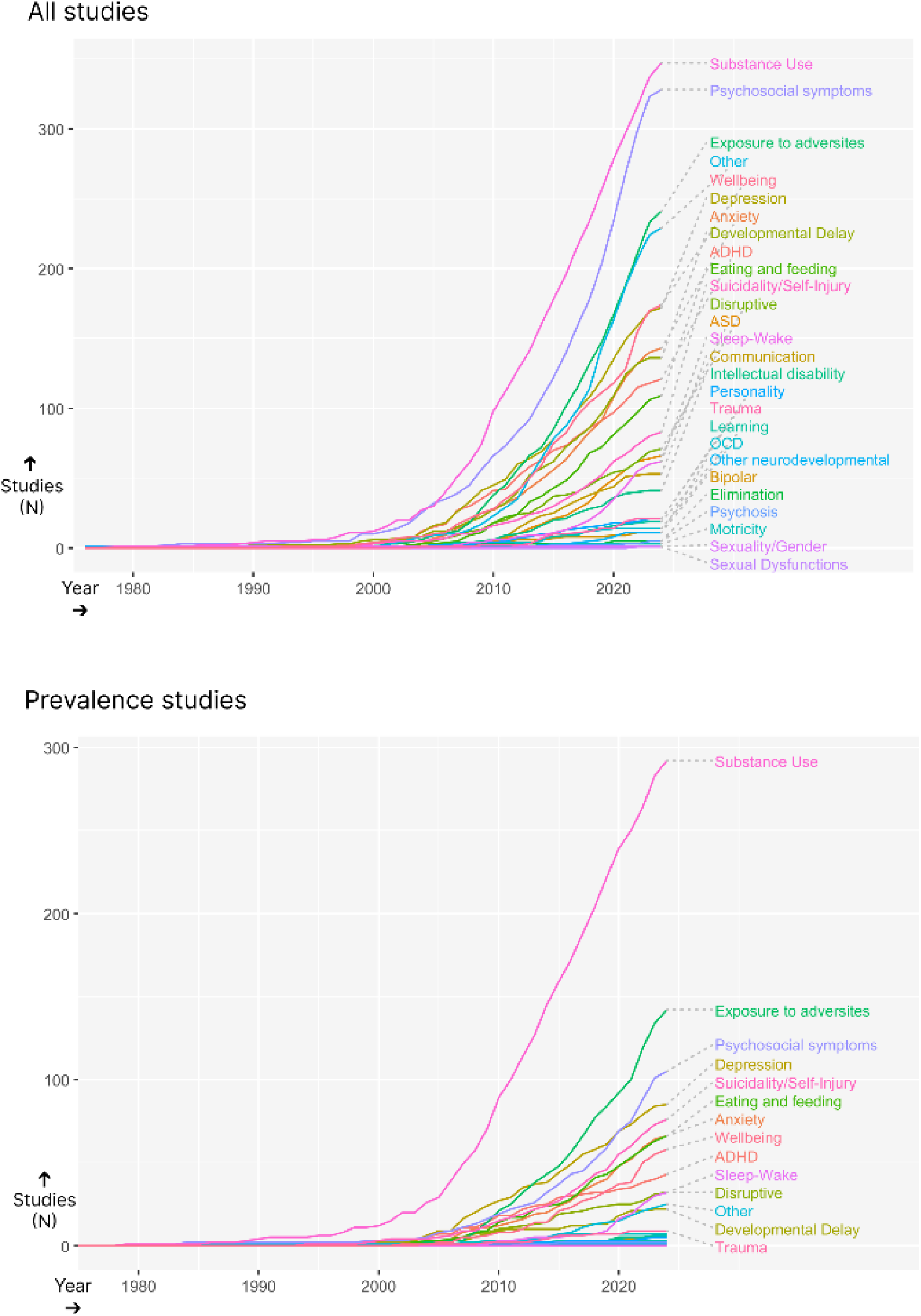
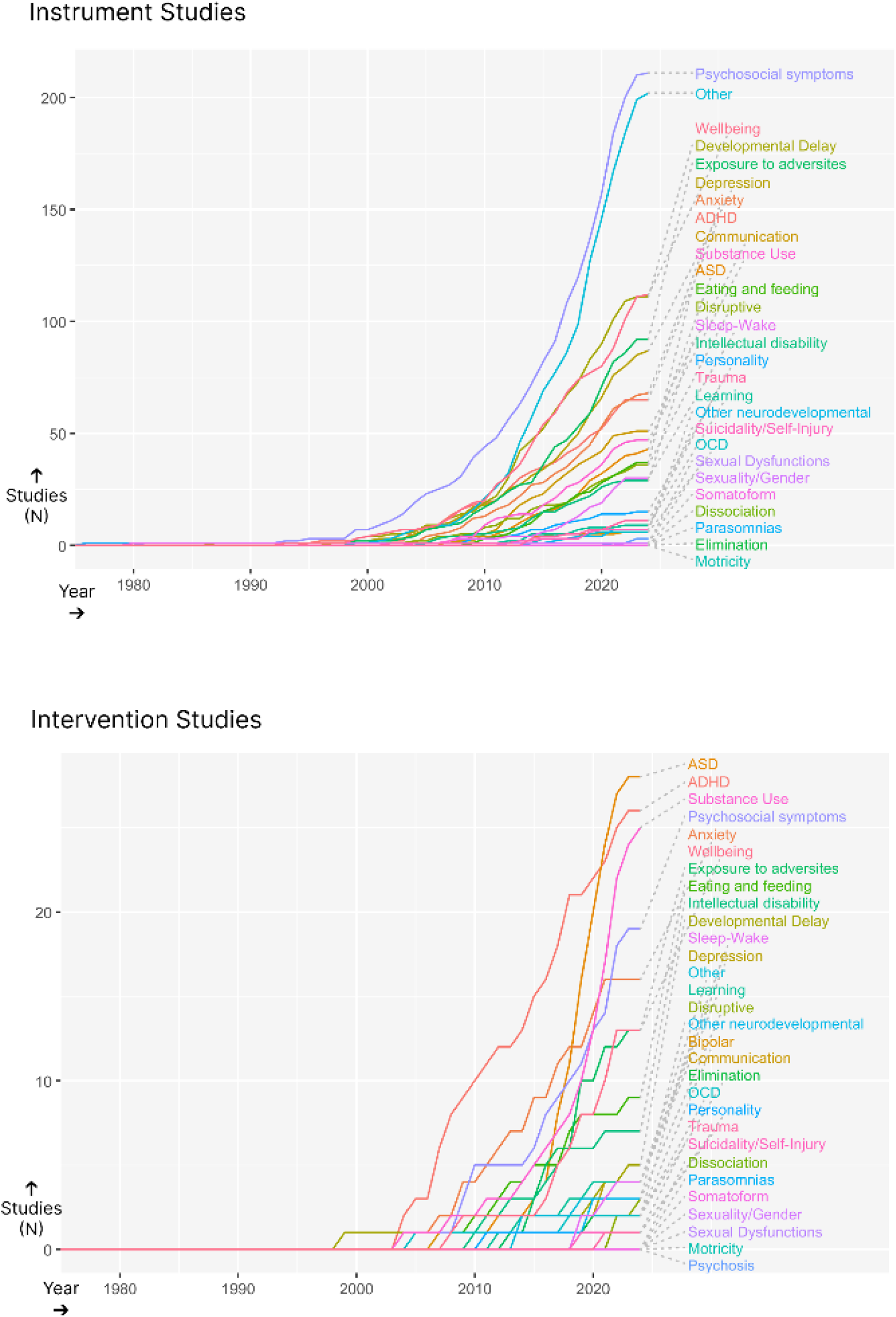
Time trend analysis and areas of concentration of scientific production on child and adolescent mental health in Brazil.

Figure 3 shows the regional distribution of prevalence surveys (see heatmap graph in Supplementary Figure 2). There is a concentration of studies in the historically wealthier and whiter southern and southeastern states, particularly São Paulo (SP) and Rio Grande do Sul (RS). This reflects a centralization of scientific production in specific universities across these regions, with studies usually representing urban populations near these academic centers.

**Figure 3.**
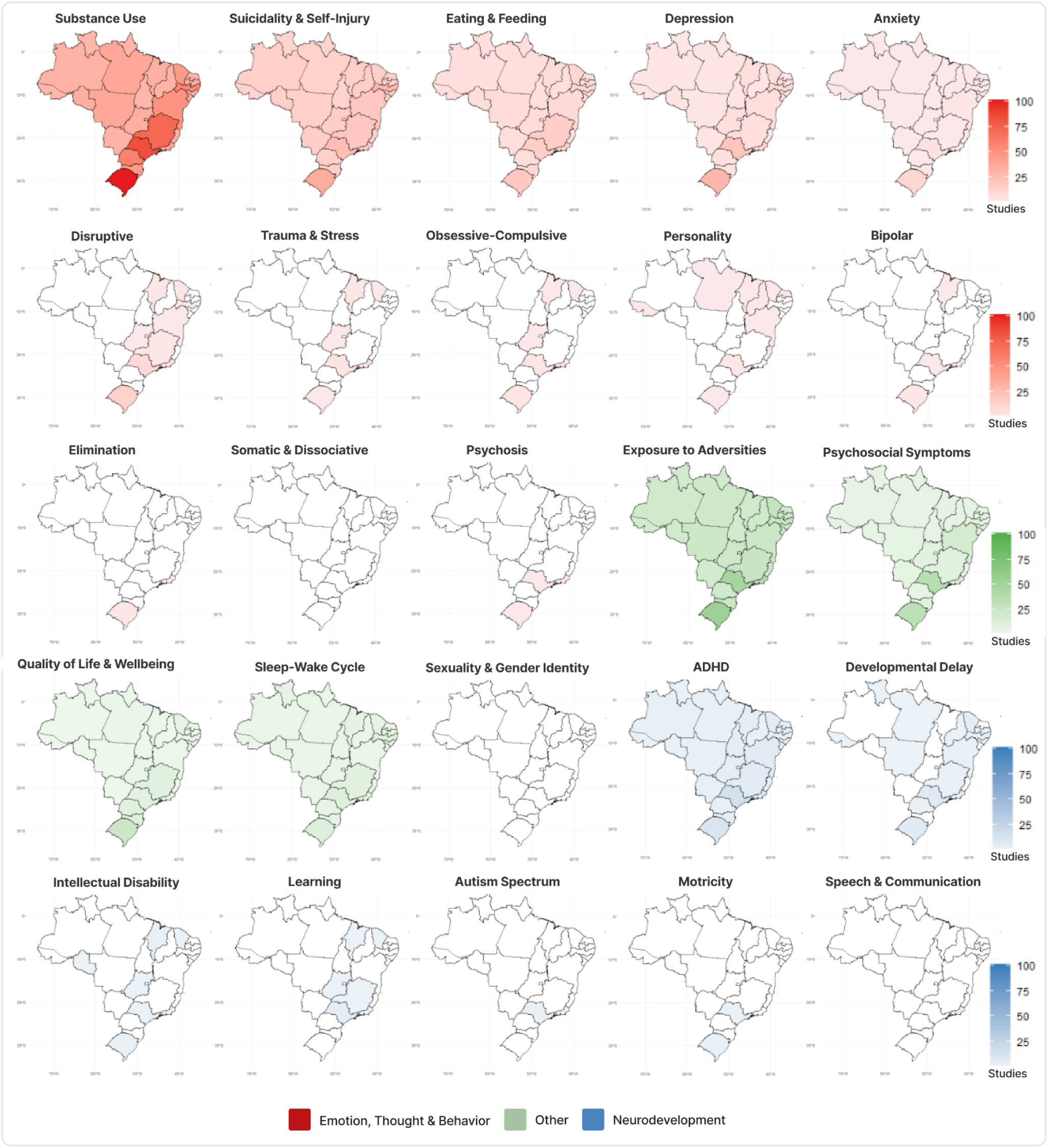
Regional analysis of represented population and conditions according to the number of studies.

### Prevalence estimates

#### Appraisal of the literature

Regarding quality parameters, 517 out of all the 734 studies (70.44%) used probabilistic samples. Most studies on the prevalence of mental disorders relied on screening questionnaires using cutoff points for establishing diagnosis or at-risk population,^41^ and only 27 studies (3.68%) employed structured clinical interviews based on standardized diagnostic criteria to confirm diagnoses. Only 69 studies (9.40%) included more than 5,000 participants, and 75 studies (10.22%) recruited participants from all five national regions. Many studies reported small samples from specific locations, with 530 studies (72.21%) recruiting participants from a single state.

There is no nationwide study with clinical interviews on mental disorders prevalence in Brazil, and best evidence is derived from high-quality studies in specific regions. There are nationwide representative surveys on several mental health-related constructs (e.g., substance use, bullying), primarily conducted by governmental entities. Official registers available from the public health system register system (DATASUS) provides nationally-representative data on self-injury notifications, hospitalizations due to psychoactive substances, and rates of violent death.

#### Findings

Table 1 selects best-available evidence on the prevalence of mental disorders (see **Supplementary File 1** for full dataset). Current prevalence of any psychiatric diagnosis ranged from 10.8% (at 12 years old in Pelotas, RS) to 19.9% (age 7 to 14, in Porto Alegre, RS, and São Paulo, SP).^42,43^

**Table 1.** Best data on the prevalence of mental disorders: selection of ten studies (N ≥ 1,000, using structured clinical interview and diagnostic manual criteria)

| Study<br>(most recent first) | Instrument / criteria | Description | City (State) Region | Year of collection | Sample size | Age groups<br>Girls percentage<br>Ethnicity | Point prevalence<br>(highest to lowest) |  |
| --- | --- | --- | --- | --- | --- | --- | --- | --- |
| Bauer et al. (2022)<sup>77</sup> | DAWBA DSM-V | Follow-up of 1993 Pelotas birth cohort<sup>94</sup> | Pelotas (RS) South | 2010 and 2015 | 4229 | 6 and 11 years (two time-points)<br>48.1%<br>Black or mixed: 38.3%<br>White: 61.7% | *(At 6 years-old)*<br>Any psychiatric disorder: 16.3%<br>Anxiety disorders: 8.9%<br>ADHD: 2.6%<br>CD/ODD: 2.6%<br>Mood disorders: 1.3% | *(At 11 years-old)*<br>Any psychiatric disorder: 13%<br>Anxiety disorders: 4.3%<br>ADHD: 4%<br>CD/ODD: 3.6%<br>Mood disorders: 3.2% |
| Álvares et al. (2021)<sup>95</sup> | MINI DSM-IV | Follow-up of 1997/1998 São Luís birth cohort<sup>96</sup> | São Luís (MA) Northeast | 2016 | 2486 | 18 - 19 years<br>52.4% | PTSD: 2.5% |  |
| Anselmi et al. (2010)<sup>42</sup> | DAWBA DSM-IV | Follow-up of 1993 Pelotas birth cohort<sup>94</sup> | Pelotas (RS) South | 2004 to 2006 | 4448 | 12 years<br>50.3%<br>Black or mixed: 28.5%<br>Asian or native: 4.7%<br>White: 66.8% | Any psychiatric disorder: 10.8%<br>ADHD: 4.1%<br>Anxiety disorder: 2.2%<br>CD: 2.2% | Major Depressive Disorder: 1.6%<br>Eating disorder: 0.1%<br>ODD: 2.1% |
| Simões et al. (2020)<sup>96</sup> | MINI DSM-IV | Follow-up of 1997/1998 São Luís birth cohort | São Luís (MA) Northeast | 2016 | 2515 | 18 - 19 years<br>52.4%<br>Black: 16.9%<br>Mixed: 62.4%<br>White: 20.7% | MDD: 11.8%<br>Bipolar disorder: 11.8% | GAD: 3.5% |
| Gallo et al. (2017)<sup>97</sup> | MINI DSM-IV | Follow-up of 1993 Pelotas birth cohort<sup>94</sup> | Pelotas (RS) South | 2011 | 3715 | 18 years<br>52.6%<br>Non-white: 23.2%<br>White: 76.8% | MDD: 6.8% |  |
| Paula et al. (2015)<sup>78</sup> | K-SADS-PL DSM-IV | Cross-sectional study | Caeté (RJ), Goianira (GO), Itaitinga (CE), Rio Preto da Eva (AM) Southeast, Center, Northeast, and North | 2010 to 2011 | 1676 | 6 - 16 years<br>51.4% | Any psychiatric disorder: 13.1%<br>Any anxiety disorder: 7.2%<br>Any ADHD: 4.5%<br>Specific phobia: 3.8%<br>Any CD/ODD: 2.3%<br>Social phobia: 2.0%<br>ODD: 1.7%<br>Separation anxiety disorder: 1.3% | Agoraphobia: 0.7%<br>OCD: 0.6%<br>CD: 0.6%<br>Any depressive disorder: 0.5%<br>GAD: 0.4%<br>PTSD: 0.3%<br>MDD: 0.2%<br>Dysthymia: 0.1% |
| Vivian et al. (2014)<sup>98</sup> | K-SADS-PL DSM-IV | Cross-sectional study | Porto Alegre (RS) South | 2009 to 2011 | 2323 | 14 - 17 years<br>46.8% | OCD: 3.22% |  |
| Salum et al. (2021)<sup>99</sup> | K-SADS-PL DSM-IV | Cross-sectional study | Porto Alegre (RS) South | 2008 to 2009 | 2457 | 10 - 17 years | Any anxiety disorder: 5.62%<br>GAD: 3.87% | Social Anxiety Disorder: 2.32%<br>Separation Anxiety Disorder: 2% |
| Fleitlich-Bilyk and Goodman (2004)<sup>79</sup> | DAWBA DSM-IV | Cross-sectional study | Taubaté (SP) Southeast | 2000 to 2001 | 1251 | 7 - 14 years<br>47% | Any psychiatric diagnosis: 12.7%<br>Any CD/ODD: 7.0%<br>Any anxiety disorder: 5.2%<br>ODD: 3.2%<br>CD: 2.2%<br>Anxiety NOS: 2.1%<br>ADHD: 1.8% | Disruptive disorder NOS: 1.5%<br>Separation anxiety disorder: 1.4%<br>Any depressive disorder: 1%<br>Specific phobia: 1.0%<br>Social phobia: 0.7%<br>PTSD: 0.1%<br>OCD: 0.1%<br>GAD: 0.4% |
| Salum et al. | DAWBA DSM-IV | Brazilian High Risk Cohort | Porto Alegre (RS) and São | 2010 to 2011 | 958* | 6 - 14 years<br>45.2% | Any disorder: 19.9%<br>Any OCD: 5.5% | Bipolar: 0.3%<br>Any PDD/Autism: 0.3% |
| (2015)<sup>43</sup> |  | Study<sup>43</sup> | Paulo (SP)<br>South and<br>Southeast |  |  | Non-white: 39.5%<br>White: 60.5%* | ADHD (combined): 2.9%<br>Panic disorder: 2.9%<br>Any CD/ODD: 2.1%<br>MDD: 1.8%<br>PTSD: 1.6%<br>GAD: 0.8%<br>Social phobia: 0.3% | Anxiety (others) :<br>0.2%<br>Agoraphobia: 0.2%<br>Eating Disorder: 0.1%<br>Tic Disorders: 3%<br>Separation Anxiety Disorder:<br>0.00%<br>Psychosis: 0% |

**Notes:** the full dataset with 734 studies on 2576 prevalence estimates can be consulted in **Supplementary File 1**.
Cruzeiro et al. (2008)<sup>100</sup> was not included here because, in spite of employing DSM-IV criteria evaluated with a structured interview, it sets two positive answers as the cut-off point for conduct disorder diagnosis, instead of three as established in the manual. \*The Brazilian High Risk Cohort study is composed by 2,512 participants, but we only extracted data for the random, community-based sample (n=958), excluding the high-risk selected sample.
**Abbreviations:** **ADHD** (Attention Deficit Hyperactivity Disorder); **CD** (Conduct Disorder); **DAWBA** (Development and Well-Being Assessment); **DSM** (Diagnostic and Statistical Manual of Mental Disorders); **GAD** (Generalized Anxiety Disorder); **K-SADS-PL** (Schedule for Affective Disorders and Schizophrenia for School Aged Children: Present and Lifetime Version); **MDD** (Major Depressive Disorder); **MINI** (Mini International Neuropsychiatric Interview); **NOS** (Not Otherwise Specified); **OCD** (Obsessive-Compulsive Disorder); **ODD** (Oppositional-Defiant Disorder); **PTSD** (Post-Traumatic Stress Disorder).

Table 2 selects nationally representative estimates of mental health-related constructs. Suicide rates per 100,000 indigenous adolescents are alarmingly high (11, age group 10 to 14, years 2010 to 2015), almost 17 times the equivalent national rate (0.652).^44^ Other studies echo these findings: municipalities with a high density of self declared indigenous population presented similar rates (11.6 for children aged 5 to 14),^45^ and reports at an indigenous reservation in Dourados (MG) found even higher rates (174 for adolescents aged 14 to 19).^46^

**Table 2.** Nationally-representative estimates of mental health-related constructs: selection of 37 studies (N ≥ 10,000, data collected since 2010, nationwide)

| Studies | Datasources | Funding | Years | Sample description | Assessment | Prevalence / Rate per 100,000 people (highest to lowest) |  |
| --- | --- | --- | --- | --- | --- | --- | --- |
| 44,61,101-104 | National Information System of Mortality (SIM)<br>The National Institute of Geography and Statistics (IBGE) | Public | 2010 - 2020 | National system notifications<br>1 to 19 years-old | Death registers<br>National population census | **Suicide mortality rates**<br>Indigenous population (10 - 14 years): **11**<br>15 - 19 years: **4.3 to 5.3**<br>10 - 19 years: **2.43 to 2.65**<br>1 - 19 years: **1.67**<br>Under 15 years: **0.59 to 0.86**<br>10 - 14 years: **0.6** | **Homicide-mortality rates**<br>1 - 19 years: **10.87**<br>**Firearm-related death rates**<br>1 - 19 years: **10.04**<br>**Motor vehicle accident death rates**<br>1 - 19 years: **5.2** |
| 47,48,60 | Notifiable Diseases Information System (Sinan)<br>The National Institute of Geography and Statistics (IBGE) | Public | 2009 - 2021 | National system notifications<br>10 to 19 years-old | Health notifications<br>National population census | **Self-inflicted injury prevalence**<br>2019 - 2021: **27.39%**<br>**Self-inflicted injury rates**<br>2019: **133.1**<br>2018: **84.2**<br>2017: **59.1**<br>2016: **32.8**<br>2015: **15.3**<br>2010: **4.7** | **Self-inflicted injury rates (occurred at school)**<br>2018: **2.75**<br>2017: **1.77**<br>2016: **0.51**<br>2015: **0.29**<br>2011: **0.09** |
| 105 | Hospital Information System of the Brazilian National Health System (SIH/SUS)<br>The National Institute of Geography and Statistics (IBGE) | Public | 2017 - 2022 | National system notifications<br>10 to 19 years-old | Hospitalization notifications (ICD-10: F10 or F19)<br>National population census | **Hospitalization rates** for mental and behavioral disorders due to alcohol or psychoactive substances<br>2022: **13.72**<br>2021: **13.18**<br>2020: **13.10** | **Hospitalization rates** for mental and behavioral disorders due to alcohol or psychoactive substances<br>2019: **17.77**<br>2018: **17.53**<br>2017: **16.18** |
| 49-56,106 | 2019 National Adolescent School-based Health Survey (PeNSE)<sup>107</sup> | Public | 2019 | 159,245 adolescents<br>13 to 17 years-old<br>Students from 6<sup>th</sup> primary to 3<sup>rd</sup> secondary grade in 4361 public and private schools, urban and | Questions adapted from the Global School Based Student Health Survey (GSHS) | **VIOLENCE**<br>**Being bullied** (past month): 40%<br>**Being bullied** twice or more (past month): 23%<br>**Physical assault by parents/guardians** (past year): 21%<br>**Sexual abuse** (lifetime): 14.6% | **SUBSTANCE USE**<br>**Alcohol use** (lifetime): 65.15%<br>**Alcohol use - being drunk** (lifetime): 47%<br>**Alcohol use** (past month): 27.7%<br>**Tobacco use** (lifetime): 24.01%<br>**E-cigarette use** (lifetime): 17.14% |
| | | | | rural, including all state capitals | | ● **Rape** (lifetime): 6.3%<br>● **Physical assault not by parents/guardian** (past year): 12.2%<br>● **Missed class due to insecurity in home-school route** (past month): 11.6%<br>● **Missed class due to insecurity at school** (past month): 10.8%<br>● **Involvement in a physical fight** (past month): 10.6%<br>● **Involvement in a fight in which someone used a melee weapon** (past month): 4.8%<br>● **Involvement in a fight in which someone used a firearm** (past month): 2.9% | ● **Illicit drug use** (lifetime): 14.3%<br>● **Illicit drug use** (past month): 4.4\$ - 5.2%<br>● **Tobacco use** (past month): 6.5%<br>**WELLBEING**<br>● **Body image dissatisfaction** (present): 23.3% |
| 56,59, 108-1 13 | 2015 National Adolescent School-based Health Survey (PeNSE)<sup>114</sup> | Public | 2015 | 118,909 adolescents 13 to 17 years-old<br>Students from 6<sup>th</sup> primary to 3<sup>rd</sup> secondary grade in 3540 public and private schools, urban and rural, including all state capitals | Questions adapted from the Global School Based Student Health Survey (GSHS) | **VIOLENCE**<br>● **Being bullied** rarely or sometimes (past month): 37.7%<br>● **Practiced bullying** (past month): 19.8% - 22% (ages 13 to 15)<br>● **Physical assault by parent/guardian** (past month): 14.5 - 16.2%<br>● **Missed class due to insecurity in home-school route** (past month): 12.8%<br>● **Missed class due to insecurity at school** (past month): 9.3%<br>● **Involvement in a fight in which someone used a melee weapon** (past month): 8.2%<br>● **Being bullied** most of the times to always (past month): 6.6%<br>● **Involvement in a fight in which someone used a firearm** (past month): 5.6%<br>● **Sexual abuse** (lifetime): 4% | **SUBSTANCE USE**<br>● **Alcohol use** (past month): 29.3%<br>● **Tobacco use** (lifetime): 18.4%<br>● **Tobacco use** (past month): 6.1% to 8%<br>**WELLBEING**<br>● **Laxative misuse / self-induced vomiting** (lifetime): 6.9% |
| 115-1 19 | 2012 National Adolescent School-based Health Survey (PeNSE)<sup>120</sup> | Public | 2012 | 109,104 adolescents<br>**Age groups**<br>● 13 to 15 years: 86%<br>● ≥ 16 years: 13,2%<br>● < 13 years: 0.8%<br>9<sup>th</sup> primary grade students from 2,482 public and private schools across all<br>state capitals and other municipalities | Questions adapted from the Global School Based Student Health Survey (GSHS) | **VIOLENCE**<br>● **Practiced bullying** (past month): 20.8%<br>● **Physical assault by parent/guardian** (past month): 11.6%<br>● **Missed class due to insecurity in home-school route** (past month): 9.1%<br>● **Involvement in a fight in which someone used a melee weapon** (past month): 8.3%<br>● **Missed class due to insecurity at school**<br>(past month): 8.0%<br>● **Involvement in a fight in which someone used a firearm** (past month): 6.9%<br>● **Being bullied** most of the times to always (past month): 7.2% | **SUBSTANCE USE**<br>● **Illicit drug use** (lifetime): 7.3%<br>● **Tobacco use** (past month): 5% |
| 121-1<br>25 | 2010 CEBRID study (Brazilian Center on Drug Information)<sup>126</sup> | Public | 2010 | 50,890 adolescents 10 to 18 years-old<br>Students from 789 public and private primary and secondary schools across all state capitals | Questions adapted from the Global School Based Student Health Survey (GSHS) | **SUBSTANCE USE**<br>● **Alcohol use** (lifetime): 59.6%<br>● **Binge drinking** (past year): 31.7% (*ages 13 to 18*)<br>● **Tobacco use** (lifetime): 25.2%<br>● **Tobacco use** (past year): 15.3%<br>● **Tobacco use** (past month): 8.6% | **SUBSTANCE USE**<br>● **Illicit drug use\*** (lifetime): 5.2%<br>● **Tranquilizers use (lifetime)**: 3.9%<br>● **Crack use** (lifetime): 0.6% |
| 127 | Study of Cardiovascular Risks in Adolescents (ERICA)<sup>128</sup> | Public | 2013 - 2014 | 85,000 adolescents 12 to 17 years-old<br>Students from 1,251 public and private schools in 273 municipalities with more than 100,000 inhabitants, including all state capitals | Items on alcohol consumption within structured questionnaire | **SUBSTANCE USE**<br>● **Alcohol use** (past month): 21,2% |  |

**Note:** the full dataset with 734 studies on 2576 prevalence estimates can be consulted in **Supplementary File 1**. \*Illicit drugs groups cannabis, cocaine/crack, and ecstasy. **Abbreviations:** Centers for Disease Control and Prevention (CDC), International Classification of Diseases - 10<sup>th</sup> edition (ICD-10), Not Applicable (NA), World Health Organization (WHO).

Self-harm notification rates among adolescents have increased markedly, from 15.3 in 2015 to 133.1 in 2019.^47^ This is partly attributed to the 2014 policy mandating health notification of self-harm, yet underreporting remains a concern.^48^ Substance use is also high, with national surveys in 2019 assessing lifetime alcohol use at 65.15% and illicit drug use at 14.3% for adolescents aged 13 to 17.^49,50^ E-cigarette consumption is an emerging concern, with 17.14% lifetime use estimated in 2019.^51^

Exposure to violence remains alarmingly high and associated with adverse mental health outcomes. Nationwide estimates from 2019 indicate substantial rates among adolescents aged 13 to 17, including bullying (40% in the past month), physical domestic violence (21% in the past year), sexual abuse (14.6% lifetime prevalence), missing school due to street insecurity (11.6% in the previous month), and involvement in firearm fights (2.9% in the previous month).^49,52-56^

A racial bias is explicit in the association of violence and mental health. A study of 1,686 adolescents in Rio de Janeiro (RJ) found that those in high-crime areas were at elevated risk for mental disorders, with black adolescents experiencing a stronger association.^57^ Another study of 973 adolescents in Salvador (BA) showed that racial discrimination, rather than race itself, was linked to higher rates of depression.^58^ Racial-based bullying was associated with increased alcohol and tobacco use among racial minorities,^59^ and self-harm rates were higher among black and brown adolescents.^60^ Firearm-related death rates reached 10,04 in 2020 (ages 10-14), predominantly composed by homicides of black and brown male youth.^61^ Notwithstanding, research focused on black population remains limited, and there are only two studies assessing quilombola mental health.^62,63^

### Assessment instruments

#### Appraisal of the literature

Replication issues were evident: 282 (30.92%) out of 912 unique instruments were reported in only one study, and only 20 (2.19%) were featured in more than five studies. Many studies employed different versions of the same instrument, only validated subscales, or created new instruments instead of existing validated ones.

Psychometric assessment represents a significant quality gap. Positive evidence of internal consistency was reported for 277 instruments (30.37%), construct validity for 112 (12.28%), structural validity for 80 (8.77%), criterion validity for 39 (4.28%), and cross-cultural validity for only 20 tools (2.19%).

Instruments available in Brazil are mostly translations of international tools: 745 instruments (81.69%) were translated from English or other foreign languages. Only 120 of these had at least one validated procedure for translation and cross-adaptation, while the others used free translation or unidentified previous translations. Often, instruments had previous Portuguese versions for adults and were directly validated for children/adolescents without prior adaptation.^64^

There were 167 instruments (18.31%) originally developed in Brazilian Portuguese, and development quality and content validity was considered adequate for only 5 and 4 of those. With few exceptions,^65,66^ development of locally-constructed instruments missed essential steps, such as involving patients and professionals in assessing item relevance/comprehensibility.

#### Findings of validation

Table 3 presents a selection of 14 instruments that were positively validated across at least five psychometric properties (see **Supplementary File 1** for all 991 tools). There are valid, reliable tools for ADHD, anxiety, ASD, or psychosocial functioning, while there are scarcer options for substance use and suicidality.

**Table 3.** Assessment instruments for mental health: selection of 14 tools (positive validation reports on at least five psychometric properties)

| INSTRUMENT | POPULATION | SETTING | RATER | ITEMS | TRANSLATION | PSYCHOMETRICS<sup>1</sup> |  |  |  |  |  |  |  |  | REFERENCES |
| --- | --- | --- | --- | --- | --- | --- | --- | --- | --- | --- | --- | --- | --- | --- | --- |
| DIAGNOSTIC INTERVIEWS FOR MENTAL DISORDERS |  |  |  |  | Translation | C C | C R | C V | I C | I R | M E | R | S V | T R | References |
| DAWBA - Development and Wellbeing Assessment | Children<br>Adolescents | Clinical<br>Community | Caregiver<br>Clinician |  | Valid<sup>79</sup> | NA | + | NA | + | + | NA | + | + | NA | 43,79,129-136 |
| ADI-R - Autism Diagnostic Interview - Revised | Children | Clinical | Caregiver |  | Valid<sup>137</sup> | NA | + | + | + | + | NA | +/- | NA | NA | 137-140 |
| SCREENING INSTRUMENTS FOR MENTAL DISORDERS |  |  |  |  | Translation | C C | C R | C V | I C | I R | M E | R | S V | T R | References |
| SNAP-IV - Swanson, Nolan and Pelham Scale | Children<br>Adolescents | Clinical<br>Community | Caregiver<br>Teacher | 18 | Valid<sup>141</sup> | +/- | + | + | + | +/- | NA | +/- | +/- | NA | 142-163 |
| CBCL - Child Behavior Checklist | Children<br>Adolescents | Clinical<br>Community | Caregiver | 118 | Valid<sup>164,165</sup> | NA | +/- | +/- | +/- | NA | ? | + | +/- | +/- | 67,131,145,152,165-202 |
| SCARED - Screen for Child Anxiety Related Emotional Disorders | Children<br>Adolescents | Clinical<br>Community | Self-report | 41 | Valid<sup>203</sup> | NA | + | + | +/- | - | NA | + | +/- | +/- | 99,142,198,203-210 |
| MFQ - Mood and Feelings Questionnaire | Adolescents | School | Caregiver | 33 | Valid<sup>211</sup> | NA | NA | + | + | + | NA | NA | + | + | 212-215 |
| SCALES ON WELLBEING |  |  |  |  | Translation | C C | C R | C V | I C | I R | M E | R | S V | T R | References |
| PedsQL 4.0 - Pediatric Quality of Life Inventory | Children<br>Adolescents | Clinical<br>Community | Caregiver<br>Self-report | 23 | Valid<sup>216</sup> | NA | NA | + | +/- | +/- | NA | + | + | NA | 216-221 |
| BMSLSS - Brief Multidimensional Students' Life Satisfaction Scale | Children<br>Adolescents | School | Self-report | 6 | Uncertain | + | NA | + | +/- | +/- | NA | NA | + | NA | 222-228 |
| RSE - Rosenberg Self-Esteem Scale | Adolescents | Clinical<br>Community | Self-report | 10 | Valid<sup>229</sup> | NA | NA | + | + | NA | NA | + | + | +/- | 5,212,229-237 |
| BESAA - Body Esteem Scale for Adolescents and Adults | Adolescents | Community | Self-report | 23 | Valid<sup>238</sup> | NA | NA | + | + | NA | NA | + | + | + | 238,239 |
| CSHQ - Children's Sleep Habit Questionnaire | Children | Clinical | Caregiver | 33 | Valid<sup>240</sup> | NA | NA | +/- | +/- | NA | NA | + | + | + | 181,240-242 |

| NEURODEVELOPMENTAL ASSESSMENT |  |  |  |  | Translation | C<br>C | C<br>R | C<br>V | I<br>C | I<br>R | M<br>E | R | S<br>V | T<br>R | References |
| --- | --- | --- | --- | --- | --- | --- | --- | --- | --- | --- | --- | --- | --- | --- | --- |
| **BSID** - Bayley Scales for Infant Development | Infant | Clinical | Health professional |  | **Valid**<sup>243</sup> | NA | + | + | +/- | NA | NA | + | + | + | 243 |
| **SSRS-BR** - Social Skills Rating System | Children | School | Self-report Teacher | 30 | **Valid**<sup>244</sup> | NA | + | + | +/- | NA | NA | NA | +/- | +/- | 233,245-251 |
| **CHEXI** - Childhood Executive Functioning Inventory | Children Adolescents | School | Health professional | 26 | **Valid**<sup>146</sup> | NA | + | +/- | + | + | NA | +/- | ? | NA | 146,148,161, 252 |

**Note:** the full dataset with 991 tools can be consulted in **Supplementary File 1**. <sup>1</sup> - “+” stands for a property with positive evidence for validation, “+/-” for mixed evidence, “-” for negative evidence, “?” for available evidence that cannot be classified as positive or negative, “NA” for non-available evidence (see *Supplementary Table 2*). **Abbreviations:** cross-cultural validity (CC), criterion validity (CR), construct validity (CV), internal consistency (IC), interrater reliability (IR), measurement error (ME), post-traumatic stress disorder (PTSD), responsiveness (R), structural validity (SV), test-retest reliability (TR).

Only two tools were cross-culturally validated to a native Brazilian language (*Karajá*), the Children Behaviour Checklist (CBCL) and the Teacher Report Form (TRF).^67^ The Everyday Discrimination Scale for Adolescents and Adults is the sole tool addressing racism.^40^

### Interventions

#### Appraisal of literature

Psychosocial interventions were reported in 139 studies (72.40%), followed by 30 pharmacological or neuromodulation studies (15.63%), and 22 lifestyle intervention studies (11.46%). School-based programs were reported in 78 studies (40.63%).

RCTs accounted for 90 studies (46.88%), followed by 65 quasi-experimental (33.85%) and 36 uncontrolled pre-post design studies (18.75%). A high risk of bias was observed in 164 studies (90.48%), including 70 RCTs. Large trials were notably lacking, with 139 studies (72.40%) involving fewer than 100 participants.

Interventions generally lacked input from individuals with lived experience, except for the single study focused on quilombolas, which employed a participatory design to develop an alcohol use intervention for adolescents.^68^

Funding sources were predominantly public, reported in 171 studies (89.06%). Considering only RCTs with at least 100 participants, public funding was present in 34 out of 35 studies.

#### Findings

**Table 4** presents a selection of interventions reported in 35 RCT with more than 100 participants (see **Supplementary File 1** for the full dataset of 192 studies). No large trial has demonstrated an effective universal school or community intervention program ready to scale in Brazil. Some interventions with null effects lacked sufficient sample power to detect outcomes in universal prevention (e.g., PERAE alcohol use prevention program with 348 participants)^69^.

**Table 4.**
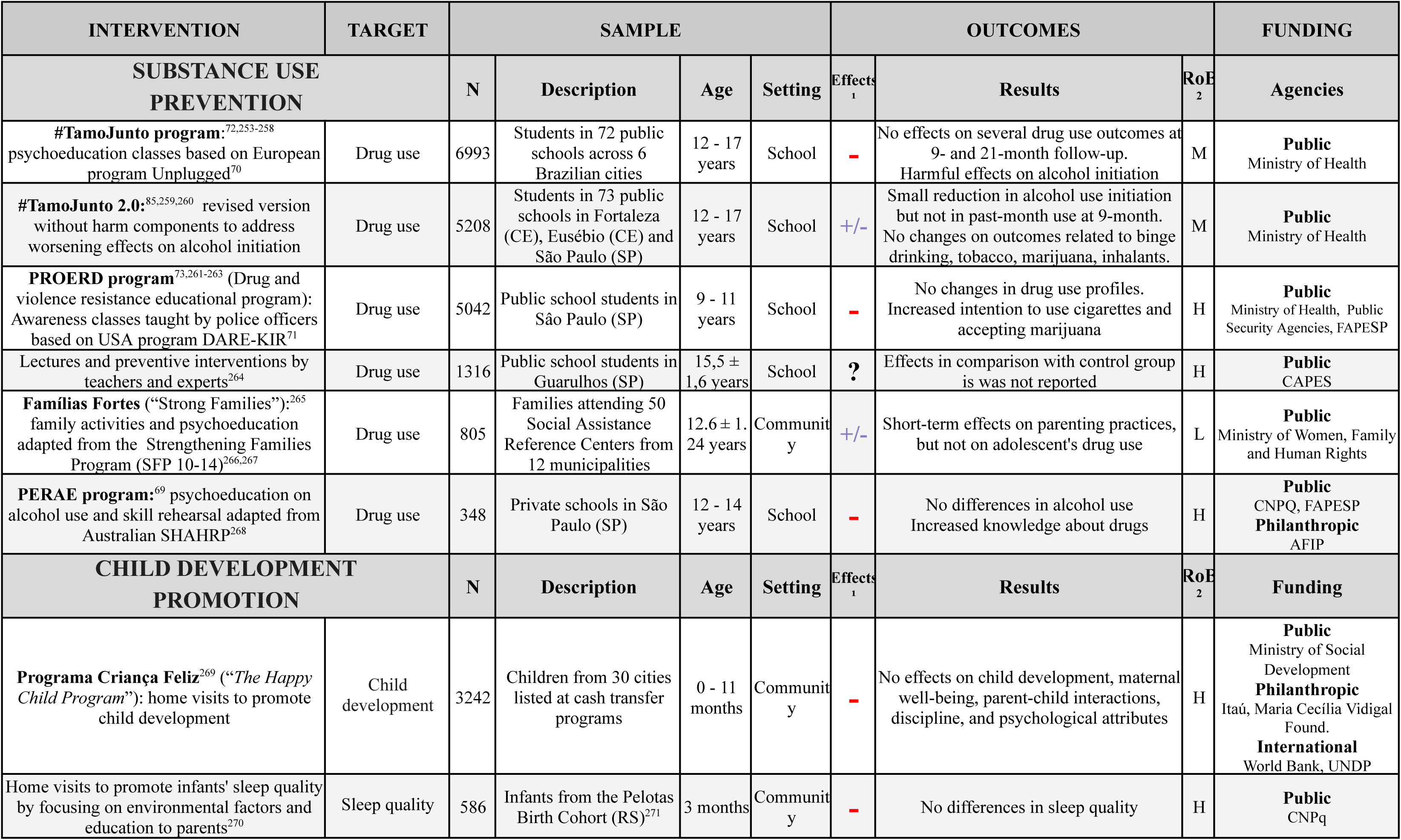
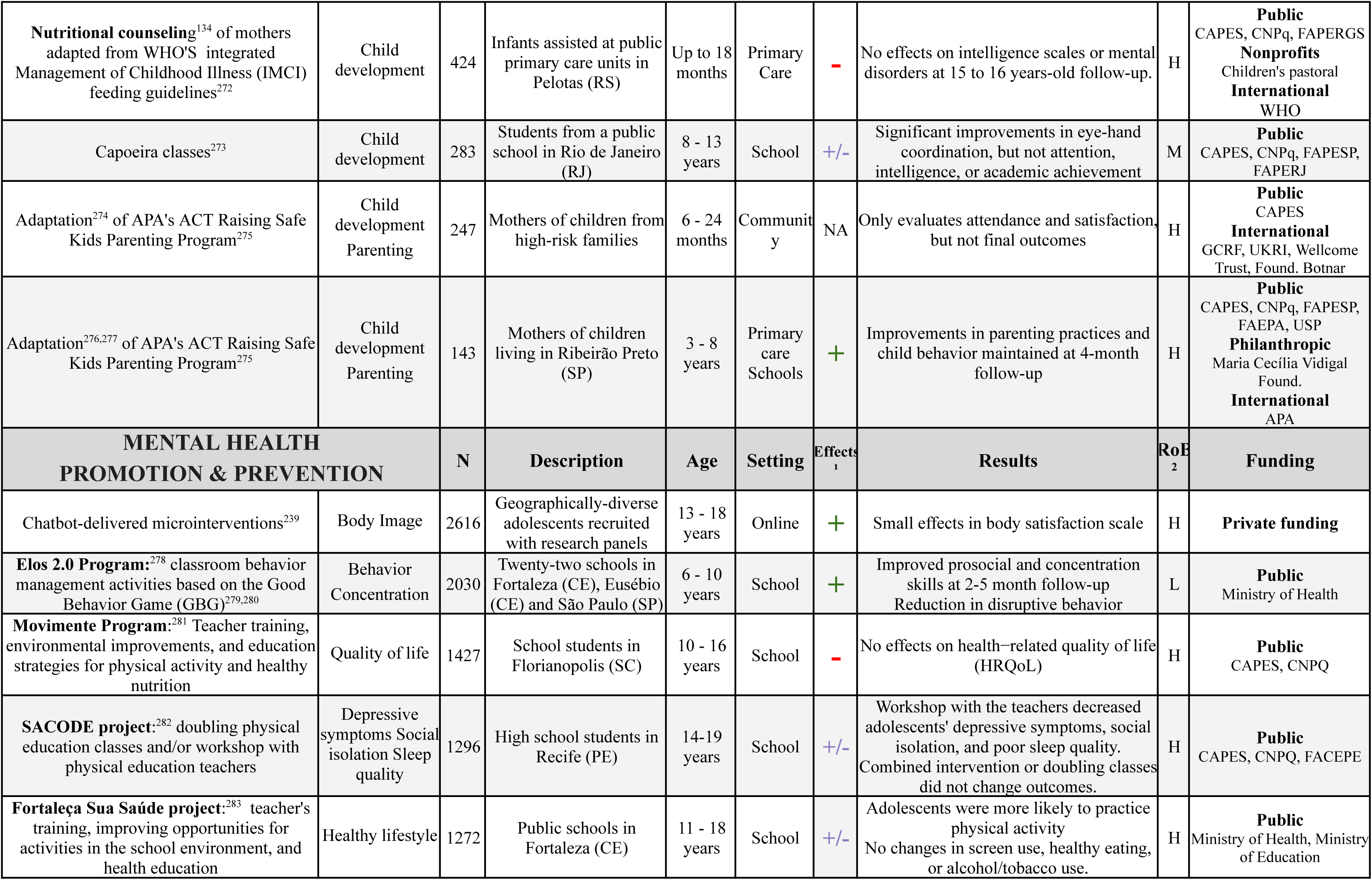
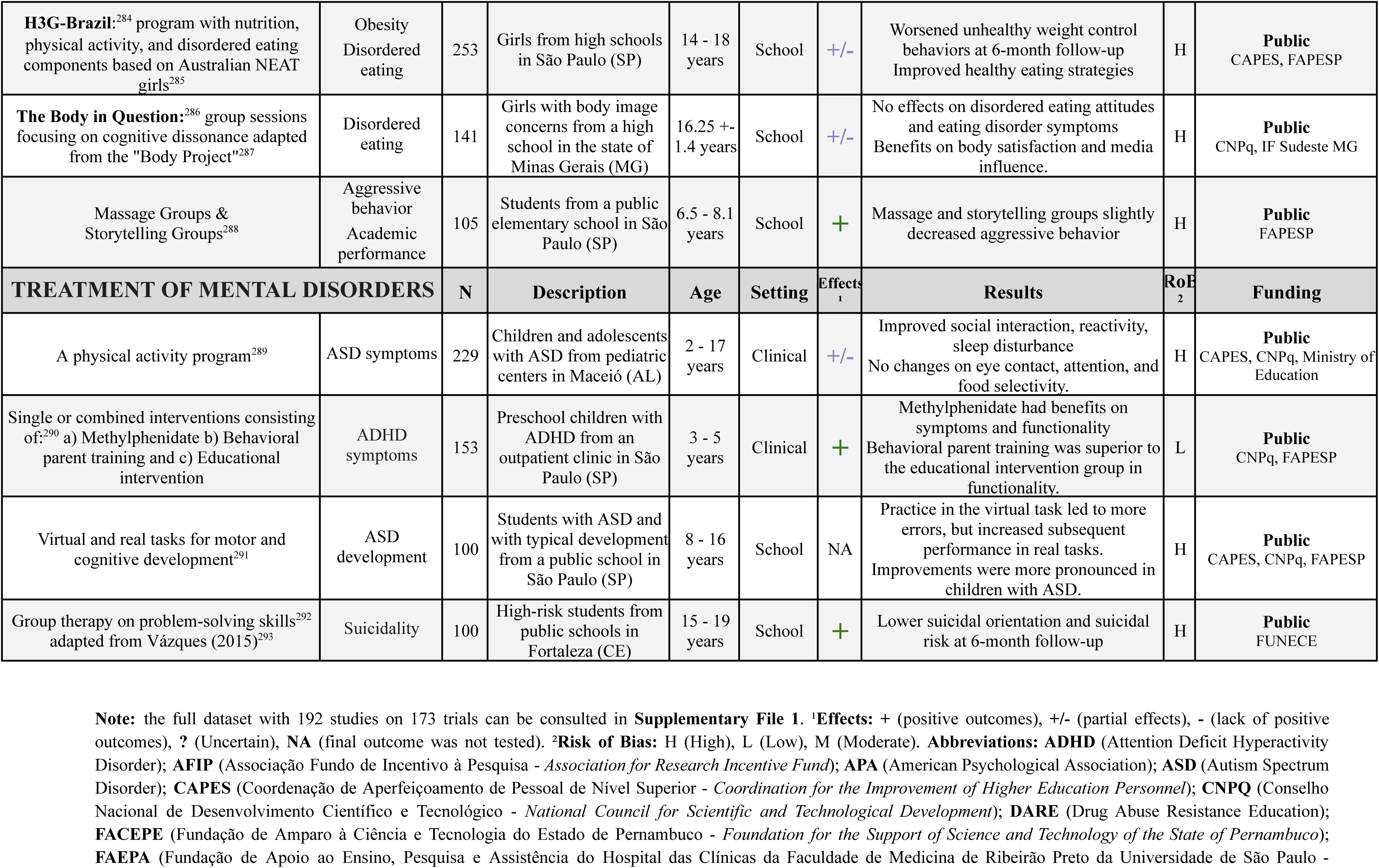
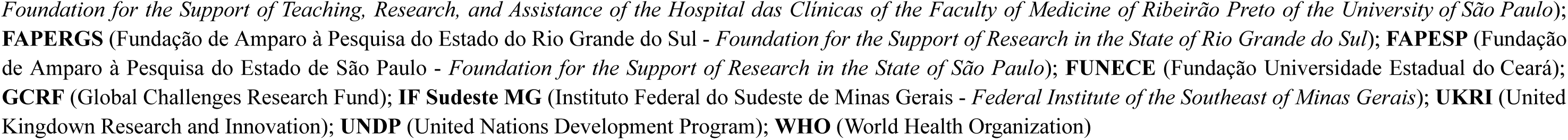
Interventions on mental health: selection of 35 studies (N > 100, randomized clinical trials)

The largest school-based programs focused on substance use prevention, including #TamoJunto, #TamoJunto 2.0, and PROERD (Drug and Violence Resistance Educational Program) which were modeled after the European Unplugged and the USA DARE programs.^70,71^ These initiatives were government-endorsed, with nationwide implementation of PROERD, yet analysis showed partial to worsening effects on drug use.^72,73^

There was a single study focusing on quilombola groups,^68^ and no interventions specifically addressed the impact of racism on mental health or targeted indigenous mental health.

## Discussion

This systematic review provides a compendium of evidence-based resources on child and adolescent mental health in Brazil. This freely-accessible dataset may facilitate the uptake of evidence-based tools among professionals, further offering a wealth of data for further scholar research and policymaking. We debate practical insights into the mental health of children and adolescents in Brazil and analyze the current state of national science in this field.

Brazil lacks a nationwide prevalence study on child and adolescent mental disorders as have been conducted in the USA and UK.^74,75^ Best available data estimate the prevalence of mental diagnoses comparable to global figures, usually around 13% for age groups from 7 to 11,^42,76-79^ yet surveys overly represent samples from south and southeast states. A most alarming finding is the high suicide rate among indigenous children and adolescents, underscoring an urgent public health issue.^44,80^ However, mental health research on indigenous youth in Brazil reflects a history of seclusion and remains very restricted.

Brazil’s burden of socio-cultural violence profoundly impacts mental health. Children and adolescents facing high rates of severe street, school, domestic, and sexual violence, increasing risks of unfavourable mental health outcomes. Systemic racism underlies many of these associations: racial discrimination, poorer living conditions, and higher exposure to violence place Black youth at elevated risk for depression, self-harm, and substance use. Notably, Brazil has a world-record rate of firearm-related adolescent deaths (10.04 per 100,000), predominantly involving homicides of young Black males.^61^ These findings add crucial evidence to the emerging debate on the mental health of Black children and adolescents in Brazil,^81^ supporting calls for intervention programs that explicitly incorporate anti-racist goals and culturally-responsive tools tailored to diverse contexts such as quilombola communities.^58,59,63^

Public funding is the primary driver of the scientific production on child and adolescent mental health. Despite growth, concerns about quality and scope persist. Many widely-used international tools lack consistent psychometric and cross-cultural validation, reflecting common challenges in non-English-speaking countries.^27^ There are no large-scale trials for anxiety, depression, stress, or bullying as reported across international literature.^82-84^ The largest school-based interventions in Brazil have adapted substance-use prevention programs from European and North American interventions, lacking efficacy. Notably, #TamoJunto 2.0 improved cross-cultural adaptation to mitigate harmful effects on alcohol initiation outcomes reported in #TamoJunto 1.0, suggesting the complexity and risk of cultural adaptation of interventions developed in high income countries.^72,85^ Improving quality also requires addressing severe replication issues. Mental health publications are numerous but fragmented and uncoordinated, leading to hundreds of duplicated efforts and isolated findings. This compendium aims to enhance access to appraised and cataloged scientific material, fostering a common language and potentially optimizing resources within the scholarly community.

This broad-scope review adopts a comprehensive search strategy, encompassing a range of databases, snowballing inclusions, expert consultations, and framed search for gaps, without restrictions of time or language. It adheres to evidence synthesis guidelines across three domains, rigorously appraising studies according to established manuals such as Cochrane and COSMIN.^35,36^ Our database synthesizes a substantial amount of data in an accessible manner for consultation, potentially representing the largest compilation effort in this field.

We also encountered limitations. The broadness of scope implies that many studies might not be located. As highlighted in COSMIN manual,^35^ capturing all data sources for instruments is challenging, and many translations and validations may exist in sources beyond the reach of our search strategy (e.g., books, developer manuals, and websites). Works with relevant information were not included due to age-related inclusion criteria, such as the gender dysphoria prevalence studies that do not discriminate information for participants under 19.^86-89^ Most prevalence and intervention studies also reported information on instruments, potentially overrepresenting the latter’s numbers in the final aggregation. We only assessed quantitative works. It is important to note that qualitative social and psychosocial research is robust in Brazil and provides culturally-sensitive insights to illuminate quantitative exploration.^90-93^

Brazilian research on child and adolescent mental health misrepresents the country’s specific struggles, as it is largely based on international perspectives that are insufficient to address local context.^15^ Brazilian specific challenges impact youth’s mental health, particularly violence, racism, and indigenous seclusion. As the primary funder of national science, public funding should direct incentives to develop culturally-sensitive assessment and scalable interventions focusing on social minorities.

## Data Availability

All data produced in the present work are contained in the manuscript

## Conflicts of interest

**Guilherme V. Polanczyk** has served as a speaker and/or consultant to Abbott, Ache, Adium, Apsen, EMS, Libbs, Medice, Takeda, and receives authorship royalties from Manole Editors.

**Luis Augusto Rohde** has received grant or research support from, served as a consultant to, and served on the speakers’ bureau of Abdi Ibrahim, Abbott, Ache, Adium, Apsen, Bial, Cellera, EMS, Knight Therapeutics, Libbs, Medice, Novartis/Sandoz, Pfizer/Upjohn/Viatris, Shire/Takeda, and Torrent in the last three years. The ADHD and Juvenile Bipolar Disorder Outpatient Programs chaired by Dr Rohde have received unrestricted educational and research support from the following pharmaceutical companies in the last three years: Novartis/Sandoz and Shire/Takeda. Dr Rohde has received authorship royalties from Oxford Press and ArtMed.

**Shekhar Saxena** is a senior advisor to McKinsey Health Institute.

**Arthur Caye** served as a consultant for Knight Therapeutics.

**Rodrigo Bressan** received institutional grants, personal fees and non-financial support from Janssen.

**All other authors** declare no conflict of interest.

All declared support occurred outside of the present work.

## Funding

Financial support has been provided by the Stavros Niarchos Foundation (SNF) as part of its Global Health Initiative (GHI) through the SNF Global Center for Child and Adolescent Mental Health at the Child Mind Institute. The funder had no role in the methodology, execution, analyses, or interpretation of the data.

## Acknowledgements

We thank Débora Renata Moura Ramos and Samanta Duarte for designing the flowchart included in this paper.

## Notes

### Summary of Updates

Enhancing the readibility of the abstract section. It was missing the line breaks and bolds. No changes were made in content.

