## Supplementary for "The science of child and adolescent mental health in Brazil: a nationwide systematic review and compendium of evidence-based resources"


**Supplementary File 1 - Full dataset** **2**

**Supplementary Figure 1 - Studies per nosological domain and year** **3**

**Supplementary Figure 2 - Prevalence studies per nosological domain and state** **4**

**Supplementary Table 1 - Preferred Reporting Items for Systematic Reviews and Meta-Analyses (PRISMA) Checklist** **5**

**Supplementary Table 2 - Search query per database** **8**

**Supplementary Table 3 - Search strategy for specific gaps** **11**

**Supplementary Table 4 - Instruments: data extracted for each instrument reported at each study** **12**

**Supplementary Table 5 - Instruments: evaluation of psychometric properties and language in summary table** **14**

**Supplementary Table 6 - Synthesis of each instrument property: coding criteria** **15**

**Supplementary Table 7- Reasons for exclusion** 16

**References** **17**

### Supplementary File 1 - Full dataset

Refer to attached file.

### Supplementary Figure 1 - Studies per nosological domain and year

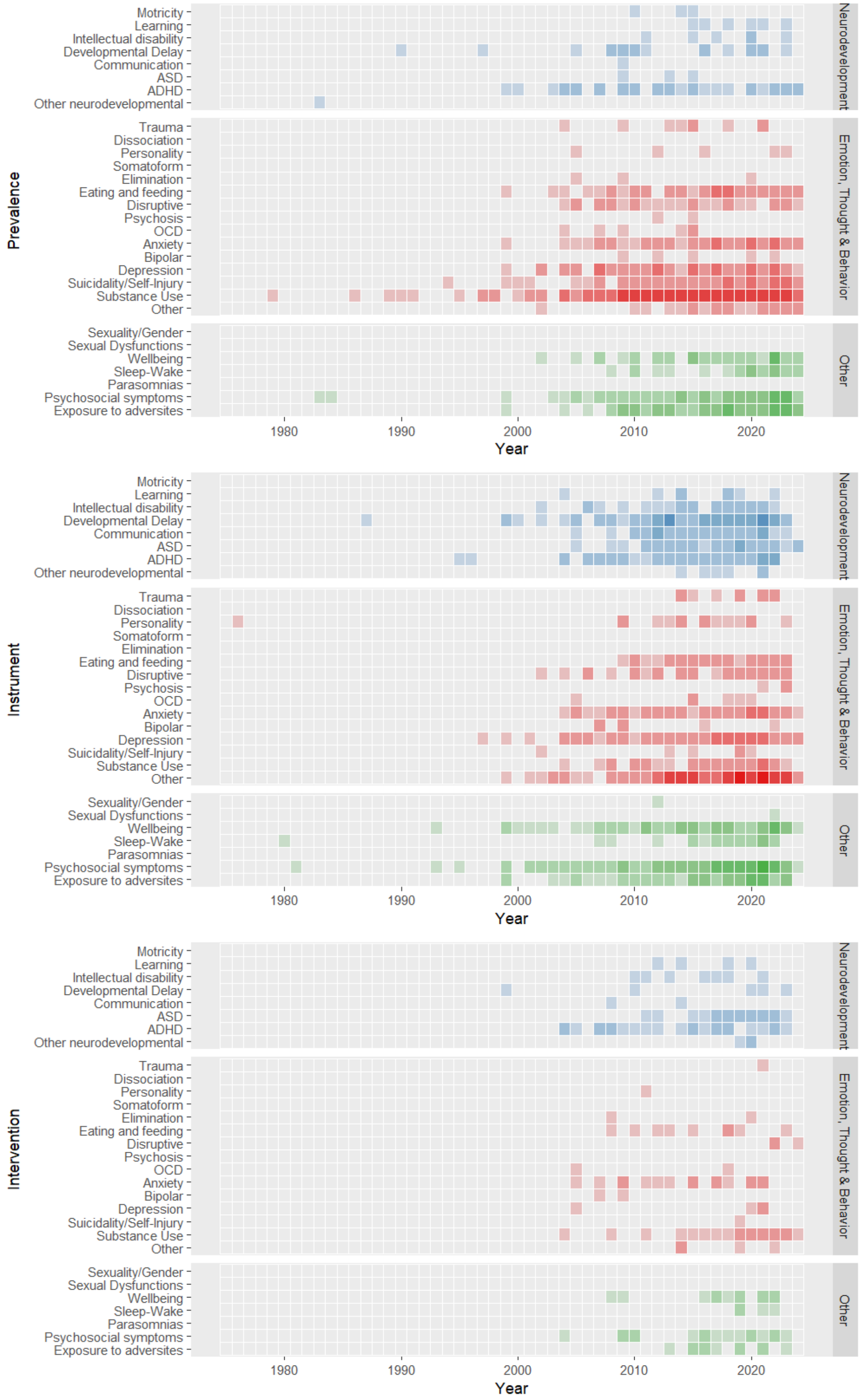

### Supplementary Figure 2 - Prevalence studies per nosological domain and state

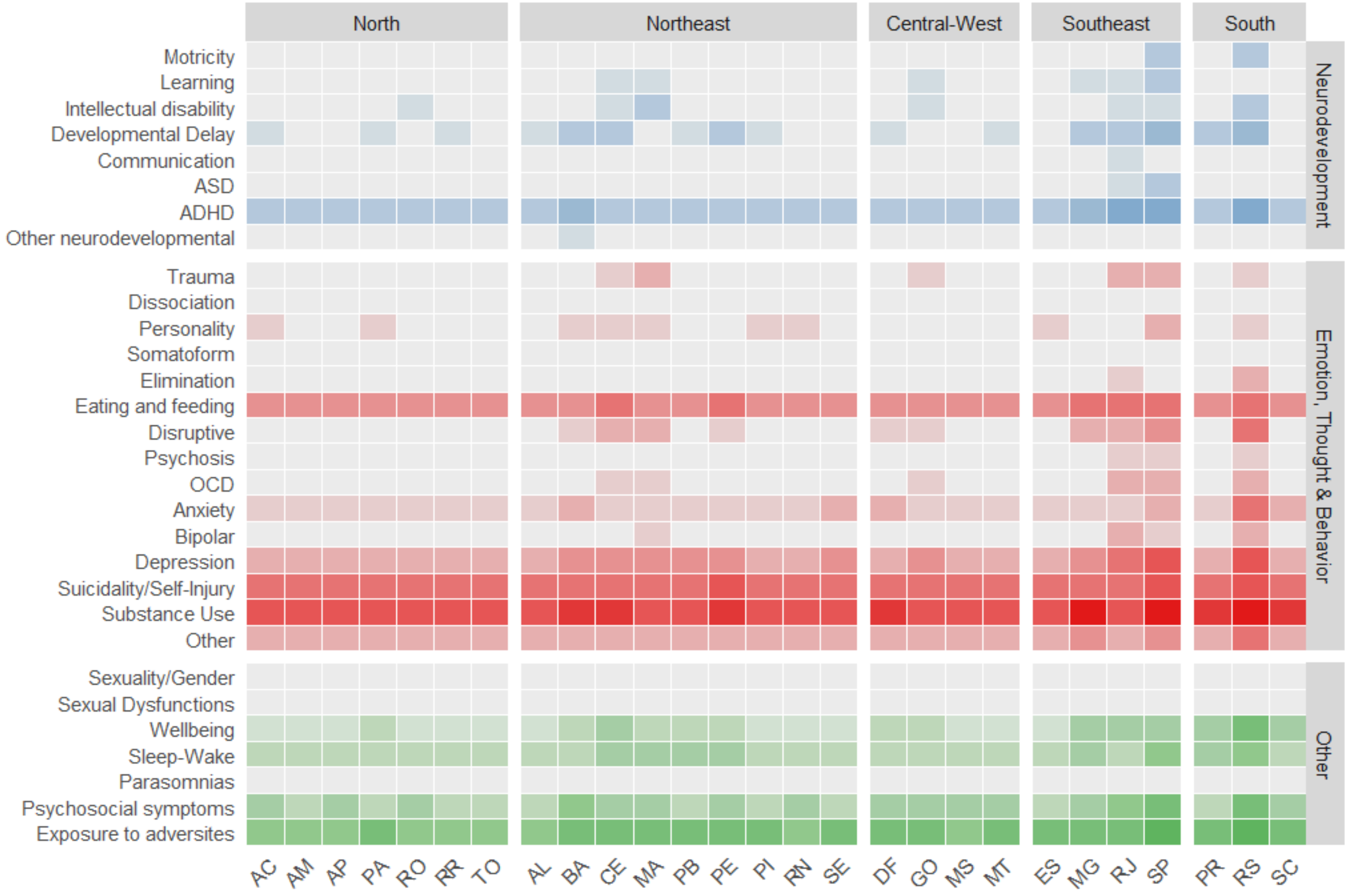

**Supplementary Table 1 - Preferred Reporting Items for Systematic Reviews and Meta-Analyses (PRISMA) Checklist**

| Section and Topic | Ite m # | Checklist item | Location where item is reported |
|---|---|---|---|
| **TITLE** | | | |
| Title | 1 | Identify the report as a systematic review. | Title |
| **ABSTRACT** | | | |
| Abstract | 2 | See the PRISMA 2020 for Abstracts checklist. | Abstract |
| **INTRODUCTION** | | | |
| Rationale | 3 | Describe the rationale for the review in the context of existing knowledge. | Introduction |
| Objectives | 4 | Provide an explicit statement of the objective(s) or question(s) the review addresses. | Introduction Methods |
| **METHODS** | | | |
| Eligibility criteria | 5 | Specify the inclusion and exclusion criteria for the review and how studies were grouped for the syntheses. | Methods |
| Information sources | 6 | Specify all databases, registers, websites, organizations, reference lists and other sources searched or consulted to identify studies. Specify the date when each source was last searched or consulted. | Methods |
| Search strategy | 7 | Present the full search strategies for all databases, registers and websites, including any filters and limits used. | Methods |
| Selection process | 8 | Specify the methods used to decide whether a study met the inclusion criteria of the review, including how many reviewers screened each record and each report retrieved, whether they worked independently, and if applicable, details of automation tools used in the process. | Methods |
| Data collection process | 9 | Specify the methods used to collect data from reports, including how many reviewers collected data from each report, whether they worked independently, any processes for obtaining or confirming data from study investigators, and if applicable, details of automation tools used in the process. | Methods |
| Data items | 10a | List and define all outcomes for which data were sought. Specify whether all results that were compatible with each outcome domain in each study were sought (e.g. for all measures, time points, analyses), and if not, the methods used to decide which results to collect. | Methods Results |
| | 10b | List and define all other variables for which data were sought (e.g. participant and intervention characteristics, funding sources). Describe any assumptions made about any missing or unclear information. | Methods |
| Study risk of bias assessment | 11 | Specify the methods used to assess risk of bias in the included studies, including details of the tool(s) used, how many reviewers assessed each study and whether they worked independently, and if applicable, details of automation tools used in the process. | Methods |

| Section and Topic | Item # | Checklist item | Location where item is reported |
|---|---|---|---|
| Effect measures | 12 | Specify for each outcome the effect measure(s) (e.g. risk ratio, mean difference) used in the synthesis or presentation of results. | Methods |
| Synthesis methods | 13a | Describe the processes used to decide which studies were eligible for each synthesis (e.g. tabulating the study intervention characteristics and comparing against the planned groups for each synthesis (item #5)). | Methods |
| | 13b | Describe any methods required to prepare the data for presentation or synthesis, such as handling of missing summary statistics, or data conversions. | Methods |
| | 13c | Describe any methods used to tabulate or visually display results of individual studies and syntheses. | Methods |
| | 13d | Describe any methods used to synthesize results and provide a rationale for the choice(s). If meta-analysis was performed, describe the model(s), method(s) to identify the presence and extent of statistical heterogeneity, and software package(s) used. | Methods |
| | 13e | Describe any methods used to explore possible causes of heterogeneity among study results (e.g. subgroup analysis, meta-regression). | NA |
| | 13f | Describe any sensitivity analyses conducted to assess robustness of the synthesized results. | NA |
| Reporting bias assessment | 14 | Describe any methods used to assess risk of bias due to missing results in a synthesis (arising from reporting biases). | NA |
| Certainty assessment | 15 | Describe any methods used to assess certainty (or confidence) in the body of evidence for an outcome. | Methods |
| **RESULTS** | | | |
| Study selection | 16a | Describe the results of the search and selection process, from the number of records identified in the search to the number of studies included in the review, ideally using a flow diagram. | Results |
| | 16b | Cite studies that might appear to meet the inclusion criteria, but which were excluded, and explain why they were excluded. | Results |
| Study characteristics | 17 | Cite each included study and present its characteristics. | Results<br>Full dataset |
| Risk of bias in studies | 18 | Present assessments of risk of bias for each included study. | Results<br>Full dataset |
| Results of individual studies | 19 | For all outcomes, present, for each study: (a) summary statistics for each group (where appropriate) and (b) an effect estimate and its precision (e.g. confidence/credible interval), ideally using structured tables or plots. | Results<br>Full dataset |
| Results of syntheses | 20a | For each synthesis, briefly summarise the characteristics and risk of bias among contributing studies. | Not applicable |
| | 20b | Present results of all statistical syntheses conducted. If meta-analysis was | Not |

| Section and Topic | Item # | Checklist item | Location where item is reported |
|---|---|---|---|
| | | done, present for each the summary estimate and its precision (e.g. confidence/credible interval) and measures of statistical heterogeneity. If comparing groups, describe the direction of the effect. | applicable |
| | 20c | Present results of all investigations of possible causes of heterogeneity among study results. | Not applicable |
| | 20d | Present results of all sensitivity analyses conducted to assess the robustness of the synthesized results. | Not applicable |
| Reporting biases | 21 | Present assessments of risk of bias due to missing results (arising from reporting biases) for each synthesis assessed. | Not applicable |
| Certainty of evidence | 22 | Present assessments of certainty (or confidence) in the body of evidence for each outcome assessed. | Full dataset |
| **DISCUSSION** | | | |
| Discussion | 23a | Provide a general interpretation of the results in the context of other evidence. | Discussion |
| | 23b | Discuss any limitations of the evidence included in the review. | Discussion |
| | 23c | Discuss any limitations of the review processes used. | Discussion |
| | 23d | Discuss implications of the results for practice, policy, and future research. | Discussion |
| **OTHER INFORMATION** | | | |
| Registration and protocol | 24a | Provide registration information for the review, including register name and registration number, or state that the review was not registered. | Methods |
| | 24b | Indicate where the review protocol can be accessed, or state that a protocol was not prepared. | Methods |
| | 24c | Describe and explain any amendments to information provided at registration or in the protocol. | Methods |
| Support | 25 | Describe sources of financial or non-financial support for the review, and the role of the funders or sponsors in the review. | Title page |
| Competing interests | 26 | Declare any competing interests of review authors. | Title page |
| Availability of data, code and other materials | 27 | Report which of the following are publicly available and where they can be found: template data collection forms; data extracted from included studies; data used for all analyses; analytic code; any other materials used in the review. | Results<br>Full dataset |

### Supplementary Table 2 - Search query per database

| **Prototypical query in English for PubMed (adapted to other database syntaxexs PsycINFO, Web of Science, Scielo, and Lilacs)** |
| --- |
| Block 1: *(“Mental disorders”[Mesh terms]) OR (Autism OR Enuresis OR Encopresis OR ADHD OR “Intellectual disability” OR “Mental retardation” OR “Oppositional Defiant Disorder” OR “Conduct Disorder” OR “Depression” OR “Bipolar” OR “Disruptive Mood Dysregulation Disorder” OR “Suicide” OR “Suicidality” OR “Self-harm” OR “Obsessive-compulsive disorder” OR “Trauma” OR “PTSD” OR “Mutism” OR “Substance abuse” OR “Cannabis” OR “Alcohol” OR “Drug abuse” OR “Anorexia” OR “Bulimia” OR “Eating disorder” OR “Borderline” OR “Personality disorder” OR “Schizophrenia” OR “Psychosis” OR "Mental health" OR "Quality of life" OR "Well-being" OR “learning disorder” OR anxiety OR phobia OR panic)*<br><br>AND<br>Block 2: *(“Brazil” or “Brazilian”)*<br><br>AND<br><br>Block 3 (only in Title): *(adolesc* OR preadolesc* OR pre-adolesc* OR child* OR boy* OR girl* OR infant* OR juvenil* OR minors OR paediatri* OR pediatri* OR pubescen* OR puberty OR school* OR student* OR teen* OR young OR youth* OR class* OR orphan* OR high-school OR "high school” OR preschool* OR pre-school*)* |
| **Prototypical query in Portuguese for Scielo (adapted to Lilacs)** |
| *(ti:((adolescente OR pré-adolescente OR criança OR menino OR menina OR bebê OR juvenil OR menores OR pediátrico OR pubescente OR puberdade OR escola OR estudante OR adolescente OR jovem OR juventude OR classe OR órfão OR "ensino médio" OR "escola secundária" OR pré-escolar))) AND ("Transtorno mental" OR Mental OR Autismo OR Enurese OR Encoprese OR TDAH OR "transtorno de déficit de atenção e hiperatividade" OR "Deficiência intelectual" OR "retardo mental" OR "transtorno opositivo desafiador" OR "transtorno de conduta" OR depressão OR bipolar OR "transtorno de desregulação de humor disruptivo" OR suicídio OR suicidabilidade OR "automutilação" OR "transtorno obsessivo-compulsivo" OR trauma OR TEPT OR mutismo OR "uso de substâncias" OR cannabis OR álcool OR "abuso de drogas" OR anorexia OR bulimia OR "transtorno alimentar" OR borderline OR "transtorno de personalidade" OR esquizofrenia OR psicose OR "saúde mental" OR "qualidade de vida" OR "bem-estar" OR “ansiedade” OR “ansioso” OR “ansiosa” “fobia” OR “fobico” OR “fobica” OR aprendizado OR cognitivo OR panico)) AND (Brasil OR Brasileiro OR Brasileira)* |
| **Adaptation to Google Scholar*** |
| Portuguese search: *(transtorno mental) AND (criança adolescente) AND (brasil brasileiro brasileira)*<br>English search: *(mental) AND (child adolescent) AND (brazil brazilian)* |
| **Adaptation to BDTD (Biblioteca Digital Brasileira de Teses e Dissertações)**** |

Abstract: *("Transtorno mental" OR "Mental" OR "Autismo" OR "Enurese" OR "Encoprese" OR "TDAH" OR "transtorno de déficit de atenção e hiperatividade" OR "Deficiência intelectual" OR "retardo mental" OR "transtorno opositivo desafiador" OR "transtorno de conduta" OR "depressão" OR "bipolar" OR "transtorno de desregulação de humor disruptivo" OR "suicídio" OR "suicidabilidade" OR "automutilação" OR "transtorno obsessivo-compulsivo" OR "trauma" OR "TEPT" OR "mutismo" OR "uso de substâncias" OR "cannabis" OR "álcool" OR "abuso de drogas" OR "anorexia" OR "bulimia" OR "transtorno alimentar" OR "borderline" OR "transtorno de personalidade" OR "esquizofrenia" OR "psicose" OR "saúde mental" OR "qualidade de vida" OR "bem-estar" OR "ansiedade" OR "fobia" OR "fobico" OR "fobica" OR "transtorno de aprendizagem" OR panico)*

AND

*Title: ("adolescente" OR "pré-adolescente" OR "criança" OR "menino" OR "menina" OR "bebê" OR "juvenil" OR "menores" OR "pediátrico" OR "pubescente" OR "puberdade" OR "escola" OR "estudante" OR "adolescente" OR "jovem" OR "juventude" OR "classe" OR "órfão" OR "ensino médio" OR "escola secundária" OR "pré-escolar")*

AND

Abstract: *("Brasil" OR "Brasileiro" OR "Brasileira")*

AND

Abstract: *("instrumento" OR "escala" OR "desfecho" OR "prevalência" OR "incidência" OR "tratamento" OR "intervenção" OR "ensaio clínico" OR "proporção" OR "porcentagem" OR "epidemiologia" OR "psicometria")*

**Note:**
***** This platform does not have clear operation of syntax and booleans, and our standard query resulted in malfunctioning returns (for instance, only returning articles related to autism and not all mental health disorders). After piloting several possible queries, we opted to limit our syntax to more comprehensive keywords, as they returned the most relevant articles featuring a broad spectrum of mental disorders and related terms. ******Although this platform contains a search mechanism that operates with Boolean operators connecting multiple fields, it faced limitations in exporting files: only 1,000 references could be exported at once, and these were not organized in a standard format recognized by reference management softwares. Therefore, we opted to manually screen the titles after searching the platform. During piloting our standard query, we found a high proportion of search outputs containing methodologies beyond the scope of this study (e.g., historical, qualitative, or reflexive approaches). To achieve an optimal balance of relevant retrievals, we tested several strategies and refined the final query as follows: terms related to children and adolescents and to mental health were restricted to the "title" field, terms related to Brazil were restricted to the "abstract" field, and an additional field was added to specify methodologies. Notably, the platform orders results based on relevance. As a confirmation, our pilot searches showed that initial pages had a higher proportion of relevant articles, with this proportion progressively decreasing in further pages.

### Supplementary Table 3 - Search strategy for specific gaps

| **Instruments with missing psychometric properties** | **Outcomes** |
|---|---|
| For the 40 assessment instruments[1] reported across the highest number of studies, we manually searched Google Scholar for studies on psychometric properties missing data on validation (e.g.: *"Strengths and Difficulties Questionnaire" + "Brazil" + "structural validation"*) | 9 studies included[1–9] |
| **Gaps on prevalence estimates** | |
| We searched Google Scholar to investigate the following areas for which few to none estimates had been found: autism spectrum disorder, gender dysphoria, and intellectual disability (e.g.: *"Autism Spectrum Disorders children adolescents brazil"*) | 0 studies included |
| **Gaps on specific topics** | |
| We searched Google Scholar for studies reporting mental health outcomes with a focus on racism, quilombola populations, and sexuality/gender (e.g. *"racism mental health children adolescents Brazil"*, "*sexuality gender assessment instrument brazil adolescent"* | 4 prevalence studies[10–13]<br>2 instrument studies[13,14]<br>1 intervention study[15] |

[1]The following instruments were included in this search: Autism Behavior Checklist (ABC); Autism Diagnostic Interview - Revised (ADI-R); Autoquestionnaire Qualité de Vie Enfant Image (AUQUEI); Baptista Depression Scale for Children and Youth (EBADEP-IJ); Barkley's Side Effects Rating Scale (SERS); Bayley Infant Neurodevelopmental Screener (BINS); Beck Anxiety Inventory (BAI); Beck Depression Inventory (BDI); Body Shape Questionnaire (BSQ); Brief Multidimensional Student's Life Satisfaction Scale (BMSLSS); Brunel Mood Scale (BRUMS); Center for Epidemiologic Studies Depression Scale (CES-D); Child Behavior Checklist (CBCL); Child Behavior Checklist 1.5 to 5 years old (CBCL/1.5-5); Child Behavior Checklist 6-18 (CBCL/6-18); Childhood Autism Rating Scale (CARS); Childhood Autism Rating Scale - Brazilian Revised Version (CARS-BR); Childhood Trauma Questionnaire (CTQ); Children Depression Inventory (CDI); Children's Sleep Habit Questionnaire (CSHQ); Development and Wellbeing Assessment (DAWBA); Eating Attitudes Test (EAT-26); Módulo de Bebidas Alcoólicas e Drogas Ilícitas da Pesquisa Nacional de Saúde do Escolar (PENSE); Olweus Bully/Victim Questionnaire (OBVQ); Pediatric Daytime Sleepiness Scale (PDSS); Pediatric Quality of Life Inventory (PedsQL 4.0) Generic Core Scale; Personal Wellbeing Index 7 (PWI-7); Personal Wellbeing Index-School Children (PWI-SC); Pittsburgh Sleep Quality Index (PSQI); Resilience Scale (RS); Rosenberg Self-Esteem Scale (RSE); Schedule for Affective Disorders and Schizophrenia for School-Age Children-Present and Lifetime Version (K-SADS-PL); Screen for Child Anxiety Related Emotional Disorders (SCARED); Social Skills Rating System (SSRS-BR) Teacher Form; Strengths and Difficulties Questionnaire (SDQ); Student's Life Satisfaction Scale (SLSS); Swanson, Nolan and Pelham Scale (SNAP-IV); Teacher Report Form (TRF); Youth Self Report (YSR).

### Supplementary Table 4 - Instruments: data extracted for each instrument reported at each study

| Topics / Procedures | Extracted information |
|---|---|
| General information | Details on the study, instrument, and sample |
| Study procedures | Whether it develops, validates and/or translates, or only applies an instruments |
| Development quality* | Methodological quality** |
| Content validity quality* | Methodological quality** |
| Instrument translation | The presence of procedures such as back-and-forth translation, independent translators, expert committee assessment, and pilot testing |
| Structural validity | Type of analysis (e.g., exploratory factor analysis, principal component analysis, or confirmatory factor analysis)<br>How many factors best accounted for how much of the variance<br>Results of statistical procedures (e.g., Root Mean Square Error of Approximation (RMSEA), Comparative Fit Index (CFI), Standardized Root Mean Residuals (SRMS))<br>Sample size and methodological quality** |
| Internal consistency | Cronbach's alpha or equivalent measures<br>Sample size and methodological quality** |
| Cross-cultural validity | Multi-group confirmatory factor analysis with measurement invariance evaluation<br>Sample size and methodological quality** |
| Inter-rater reliability | Intraclass correlation coefficient, pearson or spearman correlation, kappa score, or equivalent statistics<br>Sample size and methodological quality** |
| Test-retest reliability | Intraclass correlation coefficient, pearson or spearman correlation, kappa score, or equivalent statistics<br>Sample size and methodological quality** |
| Measurement error | Statistics on the patient's score error (e.g., standard error measurement, smallest detectable change, or limits of agreement)<br>Sample size and methodological quality** |
| Criterion validity | Performance of the instrument against a gold standard (e.g., area under the curve, sensitivity, and specificity)<br>Sample size and methodological quality** |
| Construct validity | Correlation coefficients with other instruments<br>Sample size and methodological quality** |
| Responsiveness | Detection of statistically significant differences in measures over time (e.g., after an intervention) |

| Topics / Procedures | Extracted information |
| --- | --- |
| | Sample size and methodological quality** |

**Notes:** *Only for studies reporting instrument development. **Classified as very good, adequate, doubtful, inadequate, or not applicable, following the Consensus-based Standards for the Selection of Health Measurement Instruments (COSMIN)[16] guidelines

### Supplementary Table 5 - Instruments: evaluation of psychometric properties and language in summary table

| Code | Criteria for each psychometric property evaluation |
|---|---|
| + | The property was measured and the study provides positive evidence according to standard definitions from COSMIN, except for adapted definition for responsiveness<br>● **Structural validity** with CFA with CFI >0.95/RMSEA <0.06<br>● **Internal consistency** with Cronbach's alpha > 0.7<br>● **Reliability** with Pearson's *r*, ICC or weighted Kappa ≥ 0.70<br>● **Cross-cultural validity** with no differences between group factors; reliability with ICC or Kappa >= 0.70<br>● **Measurement error** with SDC or LoA < MIC<br>● **Criterion validity** with correlation with gold standard >= 0.70<br>● **Construct validity** with correlations superior 0.10 or 0.5, depending on the hypothesis<br>● **Responsiveness** statistically significant differences between time-points |
| - | The property was measured and the study provides negative evidence according to standard definitions (e.g., Cronbach's alpha < 0.7 for internal consistency, confirmatory factor analyses with CFI <0.95/RMSEA >0.06 for structural validity) |
| +/- | Mixed outcomes that may be classified either as "+" or "-" |
| ? | Information was provided for the property, but it is insufficient to ascertain it in previous categories |
| NA | Information was not available for that property |
| **Code** | **Criteria for study language and procedures** |
| **D** | Development study (in Portuguese) |
| + | Study translates the instrument with structured procedures as back-and-forth translation, independent translators, expert committee assessment, and pilot testing |
| - | Study translated the instrument without structured procedures |
| ? | Employs a previously translated version of the instrument |

**Notes**: The summary appraisal table evaluates data from the extraction table, with each entry corresponding for each instrument procedure at each study. Refer to Consensus-based Standards for the selection of health Measurement Instruments Manual (COSMIN) for details of criteria.[16] For responsiveness, we considered sufficient ("+") evidence of validation if the instrument presents statistically significant differences in a clinical trial. This procedure was chosen because studies did not provide the area under the curve in this property, which was the original parameter in the COSMIN manual. **Abbreviations**: Confirmatory Factor Analysis (CFA), Comparative Fit Index (CFI), Intraclass Correlation Coefficient (ICC), Limits of Agreement (LoA), Minimal Important Change (MIC), Root Mean Square Error of Approximation (RMSEA), Smallest Detectable Change (SDC).

### Supplementary Table 6 - Synthesis of each instrument property: coding criteria

| Code | Criteria for psychometric properties |
|---|---|
| + | Available reports with positive validation results on that psychometric propertie |
| +/- | Available reports with both positive and negative validation results on that psychometric property |
| - | Available reports with negative validation results on that psychometric propertie |
| ? | Available reports for that psychometric property that cannot be classified as positive or negative |
| NA | No available reports for that psychometric property |
| **Code** | **Criteria for study language and procedures** |
| D | An instrument developed in Portuguese with reports on its development |
| PT-BR | Instruments originally in Portuguese without reports on its development |
| P | Instruments with a previous translation to Portuguese, without reports on its procedures |
| + | Instruments that were translated or cross-culturally adapted to Portuguese with at least one of the following procedures: back-and-forth translation, independent translators, expert committee assessment, and pilot testing |
| - | Instruments that were freely translated to Portuguese |

**Note:** synthesis table aggregates all information reported for each instrument across extracted studies

**Supplementary Table 7- Reasons for exclusion**

| Primary screening | Secondary screening (prevalence) |
|---|---|
| 1. Wrong outcome: 6958<br>2. Wrong study design: 4713<br>3. Wrong population: 4542<br>4. Wrong publication type: 349 | 1. Duplicated report: 339<br>2. Wrong study design: 376<br>3. Wrong population: 131<br>4. Wrong outcome: 212<br>5. Wrong publication type: 26 |
| Secondary screening (instruments) | Secondary screening (interventions) |
| 1. Only applies an instrument already included: 1550<br>2. Wrong study design: 437<br>3. Wrong population: 289<br>4. Wrong outcome: 235<br>5. Duplicate report: 50<br>6. Wrong publication type: 59<br>7. Not found: 10 | 1. Wrong study design: 146<br>2. Wrong outcome: 45<br>3. Wrong population: 39<br>4. Wrong publication type: 8<br>5. Duplicate report: 6 |

11 Menezes AAS, Ramos DO, Sanchez ZM, Miskolci R. Racial-based bullying and substance use: A Brazilian national cross-sectional survey among students. *J Racial Ethn Health Disparities*

2023; **10**: 1441–54.

12 Castro CT, Teixeira CSS, Cairo SMC, Lima GP, Bezerra VM, Medeiros DS. Body image and associated factors among Quilombola and non-Quilombola adolescents in a semi-arid rural area in Bahia, Brazil. *Vulnerable Child Youth Stud* 2022; **17**: 82–96.

13 Azevêdo PVB, Caixeta LF, Taveira DLR, Giglio MRP, do Rosário MC, Rohde LA. Suggestive diagnosis of attention-deficit/hyperactivity disorder in indigenous children and adolescents from the Brazilian Amazon. *Eur Child Adolesc Psychiatry* 2020; **29**: 373–84.

14 Abreu MK de A, Leme VBR, Fernandes LDM, *et al.* Escala de Discriminação Cotidiana para Adolescentes e Jovens: Adaptação e Evidências Psicométricas. *Estud Pesqui Em Psicol* 2022; **22**: 709–28.

15 Moraes-Partelli AN, Coelho MP, Santos SG, Santos IL, Cabral IE. Participation of adolescents from the Quilombola community in the creation of an educational game about alcohol consumption. *Rev Esc Enferm USP* 2022; **56**: e20210402.

16 Prinsen CAC, Mokkink LB, Bouter LM, *et al.* COSMIN guideline for systematic reviews of patient-reported outcome measures. *Qual Life Res* 2018; **27**: 1147–57.
